# Real-World Characterization of Amyloid-Related Imaging Abnormalities (ARIA) in Lecanemab Treatment at an Academic Health System

**DOI:** 10.64898/2026.01.23.26344739

**Authors:** Neguine Rezaii, John R Dickson, Jeremy N Ford, Yingnan He, Yuta Katsumi, Liliana Ramirez-Gomez, Alice Lam, Hina Shah, Steven Arnold, Marina Avetisyan, Sheena R Baratono, Alaina Bennett, Paul M. Butler, Diane Chan, Matthew DeSalvo, Mark Eldaief, Rose Gallagher, Stephen Gomperts, Anna Goodheart, Mark Albers, Raymond Y Huang, Isaiah Kletenik, Sophal Lam, Michael McCormick, Samantha Milano, Nathan Praschan, Saurabh Rohatgi, Elmira Hasanzadeh, Javier Romero, Alberto Serrano Pozo, Abigail Shaughnessey, Andrew Stern, Jason You, Geoffrey Young, Jasmeer Chhatwal, Kirk R Daffner, Michael G Erkkinen, Seth Gale, Teresa Gomez-Isla, Gad A Marshall, Scott M McGinnis, Dennis Selkoe, Hyun-Sik Yang, Wai-Ying Wendy Yau, Sudeshna Das, Bradford C Dickerson

## Abstract

**Background:** The benefits of amyloid-β monoclonal antibodies for Alzheimer’s disease are tempered by the risk of amyloid-related imaging abnormalities (ARIA). Detailed real-world characterization of ARIA, including incidence, timing, radiologic severity and localization, natural history, and risk factors, is essential to optimize treatment safety. This study provides a comprehensive description of ARIA in a large real-world cohort of patients receiving lecanemab.

**Methods:** In this retrospective cohort study from the Mass General Brigham Alzheimer’s Therapeutic Program, we analyzed data from 468 patients with early Alzheimer’s disease who were at least 90 days from their first lecanemab infusion, including those whose treatment was modified or discontinued. ARIA was monitored using a standardized MRI protocol. High-dimensional analysis of baseline clinical, laboratory, and biomarker variables was performed using univariate correlations and Cox proportional hazards models with data-driven cutpoints.

**Findings:** The overall incidence of any ARIA was 25.2%, with ARIA-H occurring in 22.4% of patients and ARIA-E in 12.2%. Symptoms developed in 4.5% of all patients. Both subtypes demonstrated significant occipital involvement, with ARIA-H showing additional frontotemporal predominance. ARIA-H was typically mild and persistent with rare radiologic resolution, whereas ARIA-E was transient, resolving with a mean time to resolution of 75.6 days. Key baseline predictors of ARIA-H included CSF Aβ42 ≤683.8 pg/mL (HR 6.39), ≥1 microhemorrhage (HR 2.55), and at least one APOE ε4 allele (HR 1.77). ARIA-E was predicted by diastolic blood pressure >75 mmHg (HR 3.12), serum chloride >105 mmol/L (HR 2.79), at least one APOE ε4 allele (HR 2.51), and serum sodium >141 mmol/L (HR 1.70). ARIA-Mixed was associated with elevated serum chloride >105 mmol/L (HR 3.92), at least one APOE ε4 allele (HR 2.87), and diastolic blood pressure >75 mmHg (HR 2.60).

**Interpretation:** This comprehensive real-world characterization of ARIA, together with the identification of novel modifiable risk factors for ARIA-E, including elevated diastolic blood pressure and high-normal serum electrolytes, enables personalized risk assessment and tailored monitoring, thereby advancing the safe implementation of disease-modifying Alzheimer’s therapies.

**Funding:** Funding for this project was provided by the Mass General Neuroscience Transformative Scholar Award.

## Introduction

Anti-amyloid-β (Aβ) monoclonal antibodies, including lecanemab and donanemab, represent a landmark therapeutic advance by slowing clinical decline in Alzheimer’s disease (AD).^1,2^ This benefit, however, is counterbalanced by the risk of amyloid-related imaging abnormalities (ARIA), which encompass hemorrhage (ARIA-H) and vasogenic edema (ARIA-E).^3,4^ Real-world data characterizing ARIA outside of clinical trials remain sparse, which may cause patient hesitation even among those likely to benefit safely, and may result in adverse events for patients whose safety profile was incompletely assessed. A clearer understanding of ARIA incidence, timing, natural history, and risk factors is essential to optimize treatment safety, guide personalized monitoring, and support informed shared decision-making.

ARIA-H includes cerebral microhemorrhages and superficial siderosis, reflecting deposition of hemosiderin,^5,6^ which are best detected on T2*-weighted MRI sequences, including gradient-recalled echo (GRE) or susceptibility-weighted imaging (SWI).^7^ ARIA-E reflects vasogenic edema or sulcal effusions resulting from extravasation of protein-rich fluid into the parenchymal interstitium or leptomeningeal/subpial spaces.^3,8^ On MRI, ARIA-E manifests as a hyperintense signal in the cortex, subcortical white matter, or sulci on T2-weighted and fluid-attenuated inversion recovery (FLAIR) sequences.

The risk of developing ARIA is highest early in the course of anti-amyloid therapy.^9,10^ ARIA-H occurs in 0.5–28.4% of treated patients, whereas ARIA-E incidence ranges from 0.9–40.6%.^11^ In the Phase 3 Clarity AD trial of lecanemab, ARIA-H occurred in 17.3%, and ARIA-E was observed in 12.6% of treated patients.^1^ Established risk factors for ARIA include the presence of baseline cerebral microhemorrhages and *APOE* ε4 carrier status, with the highest risk seen in *APOE* ε4 homozygotes.^6,10,12–14^ Medical history factors that further predispose to ARIA include pre-existing autoimmune or inflammatory diseases, seizures, and cerebrovascular disease.^7^ These factors have informed appropriate use guidelines for selecting patients eligible to receive anti-Aβ monoclonal antibody therapy. The risk of ARIA-H is also increased by the presence of baseline cerebral microhemorrhages.^15–18^ Additionally, in a study of gantenerumab, lower CSF Aβ42 increased the risk of ARIA-E.^19^

Despite this knowledge, critical gaps remain in translating these findings to real-world care. The natural history, including the timing of onset and the trajectory of severity and resolution for each ARIA subtype, is not fully characterized. Furthermore, assessment of additional clinical, laboratory, and biomarker variables could identify novel, actionable risk factors and refine predictive models.

To address these knowledge gaps, we conducted a comprehensive, real-world analysis of patients receiving lecanemab within the Mass General Brigham (MGB) Alzheimer’s Therapeutic Program. Employing structured monitoring protocols and additional electronic health record data, this analysis aims to: (1) describe the incidence, timing, localization, cumulative risk profiles, and natural history of ARIA-E and ARIA-H; (2) conduct a screen for baseline predictors across multiple domains; and (3) determine data-driven clinical thresholds for actionable risk stratification.

## Methods

### Mass General Brigham Alzheimer’s Therapeutic Program

This retrospective cohort study was conducted under MGB Institutional Review Board-approved protocols (2005P000860 and 2025P001042). The study used clinical information recorded in the electronic health record to investigate ARIA in patients treated with anti-amyloid therapy. The participants of the study were patients evaluated in the MGB Alzheimer Therapeutics Program (ATP), a specialized clinical program dedicated to the evaluation and management of patients with early Alzheimer’s disease receiving anti-amyloid therapy. The inclusion criteria were referral to the MGB ATP, being at least 90 days into the treatment course, and having at least one MRI performed for ARIA monitoring. Among patients meeting these inclusion criteria, there were no additional exclusion criteria.

Patients referred to the MGB ATP undergo a clinical evaluation for eligibility for anti-amyloid therapy, guided by the eligibility criteria in the phase 3 lecanemab clinical trial^1^ and appropriate use criteria.^20^ A clinical diagnosis of mild cognitive impairment or mild dementia was required for eligibility in the program. This diagnosis was supported by standardized evaluations consisting of Mini-Mental State Examination (MMSE)^21^ or Montreal Cognitive Assessment (MoCA),^22^ Functional Activities Questionnaire (FAQ),^23^ and Quick Dementia Rating System (QDRS).^24^ The etiology of cognitive impairment was determined by cerebrospinal fluid Alzheimer’s disease biomarker positivity or amyloid positron emission tomography (PET) positivity. Some but not all cases in this cohort with amyloid PET had amyloid PET Centiloids generated in their clinical record using MIM Neuro software. APOE genotyping was required for ARIA risk stratification. For patients with an APOE genotype other than ε4/ε4, magnetic resonance imaging (MRI) with a GRE sequence showing >4 microhemorrhages was exclusionary for treatment. Given the higher risk of ARIA in patients with an APOE genotype of ε4/ε4, more stringent MRI criteria were applied to patients with this APOE genotype, with MRI GRE sequencing showing >1 microhemorrhage or susceptibility weighted imaging (SWI) showing >3 microhemorrhages being exclusionary. For all patients, the presence of superficial siderosis on MRI was exclusionary.

Eligible patients were required to sign a clinical informed consent for anti-amyloid therapy to commence. As lecanemab was the first FDA approved and clinically available agent, we have a substantially larger cohort treated with lecanemab compared to other anti-amyloid therapies at the time of this analysis. Infusions were provided at Mass General Brigham infusion centers. Lecanemab is administered at a dose of 10 mg/kg intravenously (IV) every two weeks.

### Implementation of MRI monitoring protocol in the MGB ATP

MRI surveillance for ARIA was performed using a dedicated protocol at baseline and during treatment at specific schedules. ARIA-H was detected on GRE and SWI sequences, and ARIA-E on FLAIR sequences. Our analysis here focuses on microhemorrhage counts obtained from the GRE sequence. Severity was graded using established radiologic criteria (for detailed MRI protocols, monitoring schedules, and severity grading definitions, see Supplementary Material).^25^

### Electronic health record data extraction and data harmonization

Clinical data were extracted retrospectively from electronic health records for all individuals who initiated lecanemab treatment. An AI-assisted natural language processing pipeline was developed to extract structured information from clinical notes and was supplemented by a comprehensive manual review to address missing values. Variables analyzed included three broad categories of patient characteristics, AD-specific biomarkers, and imaging/laboratory/vital sign measures. MoCA scores were converted to MMSE equivalents,^26^ and cerebrospinal fluid biomarker values were harmonized across two different assay platforms (see Supplementary Material for complete variable list with data availability and detailed methods on data preprocessing and harmonization).

### Analysis of Cumulative Incidence with Severity Stratification

To generate cumulative incidence curves stratified by radiologic severity, we conducted a time-to-event analysis using the Kaplan-Meier method with infusion number as the time metric. For patients who developed ARIA, the event time was defined as the infusion number at which the abnormality was first radiologically confirmed, and they were assigned a severity grade (mild, moderate, or severe) based on standardized criteria from their initial detection scan. Patients without ARIA were right-censored at their last completed infusion. Individual ARIA events were then plotted as points overlaid on the respective subtype’s cumulative incidence curve, with color coding corresponding to the initial severity grade. A log-rank test was performed to compare the time-to-event distributions between ARIA-E and ARIA-H.^27^

### Determining the regional prevalence of ARIA

Regional counts for ARIA-H and ARIA-E were aggregated, and the percentage of total lesions within each region was calculated. Statistical significance was determined by comparing observed lesion counts against an expected uniform spatial distribution using a one-tailed Z-test approximation of a Poisson test. Resulting p-values underwent False Discovery Rate (FDR) correction for multiple comparisons at alpha=0.05. Prevalence of ARIA-H and ARIA-E in each of the cortical lobes was then projected to *fsaverage* surface space for visualization purposes using the lobar mapping described in Klein and Tourville (2012).^28^

### Characterizing the natural history of ARIA

To characterize the natural history of ARIA, we employed a joint longitudinal-survival modeling approach. This statistical framework simultaneously analyzes two interrelated processes: the temporal evolution of ARIA severity (longitudinal component) and the time to radiologic resolution (survival component). For each patient, the date of first ARIA detection was established as time zero. All subsequent serial MRI assessments were used to construct individual severity trajectories. Resolution was defined as the first follow-up scan demonstrating complete radiologic clearance. The joint model accounts for informative censoring, where patients with persistent or severe ARIA may have differential follow-up patterns and formally assesses the relationship between an individual’s evolving severity and their subsequent probability of resolution.

### Univariate Feature Selection and Multiple Testing Correction

We examined the association of a wide range of variables with ARIA-E and ARIA-H. For each continuous variable, we calculated point-biserial correlations to assess its relationship with each ARIA type. For categorical variables, we computed Cramér’s V to measure association strength.^29^ To address the multiple comparisons inherent in our high-dimensional dataset, we applied FDR correction.

### Time to event analysis for ARIA correlates

We employed time-to-event analysis to evaluate risk factors for ARIA during lecanemab treatment. The observational period spanned from treatment initiation until the first occurrence of ARIA-E or ARIA-H, treatment discontinuation, or study completion. Patients without ARIA events were right censored at their last completed infusion.

Continuous baseline variables were evaluated for association with time-to-ARIA using a data-driven optimal cutpoint approach. For each variable, all possible dichotomization points were considered while ensuring that each resulting subgroup contained at least 15% of the analyzable population. The cutpoint that maximized the log-rank test statistics were selected as the optimal threshold. Log-rank p-values from this maximization procedure were collected for all tested continuous predictors. To control the false discovery rate across the full set of continuous variables examined, the Benjamini–Hochberg adjustment was applied globally. Variables with p_FDR_< 0.05 were declared statistically significant.

For each identified threshold, a Cox proportional hazards model was fitted using the binary high-risk versus low-risk grouping to estimate hazard ratios with 95% confidence intervals. Kaplan–Meier estimates were used to visualize cumulative incidence and to calculate absolute event rates in each risk stratum. When the direction of risk was unexpected (e.g., lower values conferring higher risk), this was explicitly noted and confirmed by both log-rank and Cox modeling results.

## Results

### MGB-ATP lecanemab sample characteristics

The cohort for this analysis consisted of 468 early-stage AD patients who had reached at least 90 days post-first infusion. Details on the demographic and clinical characteristics of this sample are presented in Table 1. The average number of infusions was 21.0 (range 1-48) (Figure 1A).

**Table 1.** Demographic and clinical information of the patients in this sample.

|  |  |  |
| --- | --- | --- |
| Variable | | Mean ± SD or % |
| **Age** | | 72.2 ± 7.8 |
| **Sex** | Female | 57.5% |
|  | Male | 42.5% |
| **Race** | White | 94.2% |
|  | Black | 1.3% |
|  | Asian | 1.5% |
|  | Other | 3.0% |
| **Ethnicity** | Non-Hispanic | 89.1% |
|  | Hispanic | 2.6% |
|  | Unavailable | 8.3% |
| **Handedness** | Right | 84.4% |
|  | Left | 11.3% |
|  | Other | 4.3% |
| **APOE e4 status** | Non-carrier | 36.8% |
|  | Heterozygote | 51.5% |
|  | Homozygote | 11.8% |
| **FAQ Total Score** | | 9.2 ± 6.3 |
| **QDRS Total Score** | | 4.8 ± 2.7 |
| **MMSE Total Score** | | 25.1 ± 3.1 |
| **Smoking** | Never | 58.1% |
|  | Former | 37.6% |
|  | Current | 3.0% |
|  | Unknown | 1.3% |

**Figure 1.**
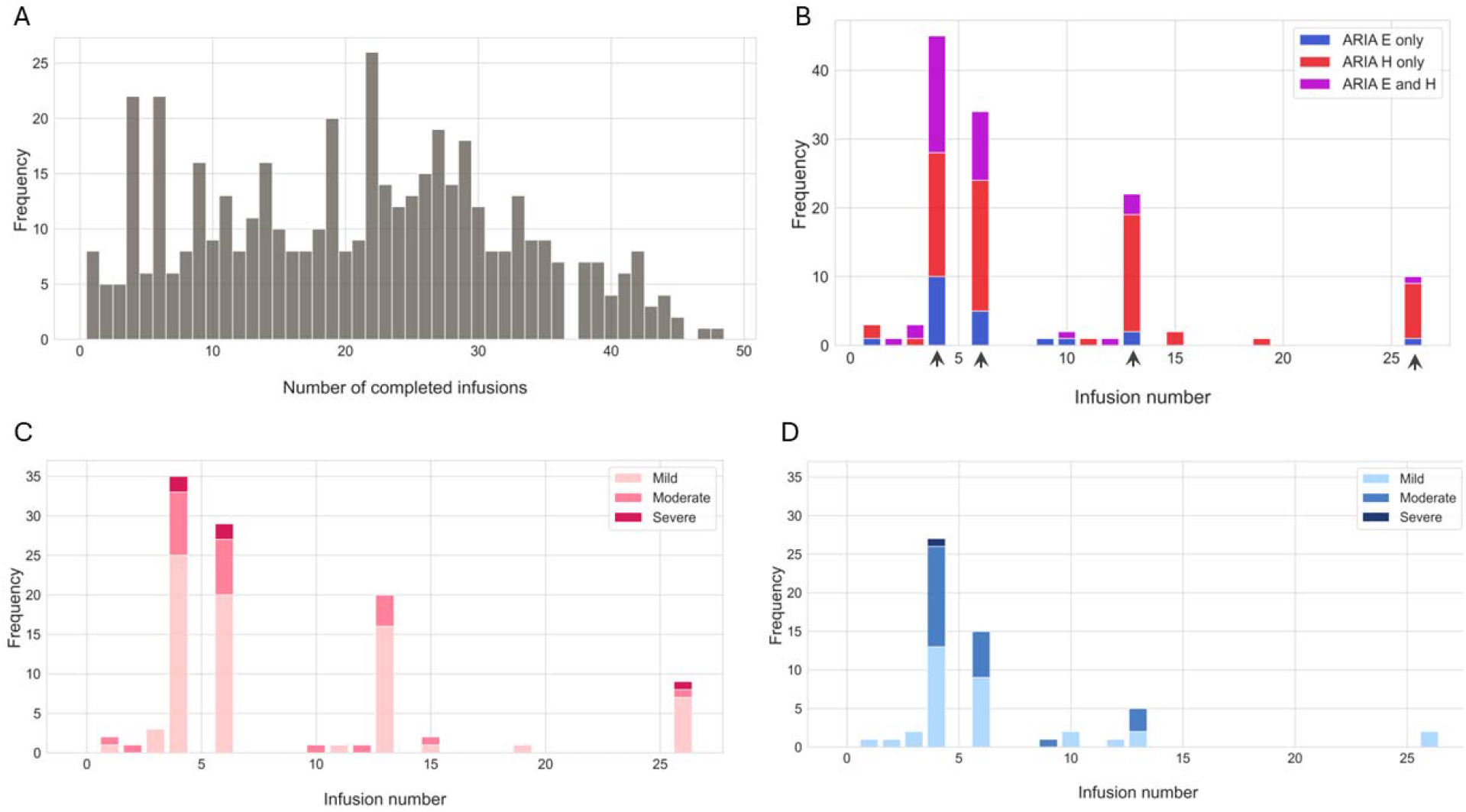
**A**. The distribution of the number of completed infusions. **B**. The time distribution of ARIA color-coded by type. Arrowheads show the time for mandatory surveillance MRIs (an additional MRI was subsequently added prior to infusion 3). **C**. Frequency of ARIA-H events by severity and infusion number. **D**. Frequency of ARIA-E events by severity and infusion number.

### ARIA incidence

Within this cohort, the overall incidence of any ARIA was 25.2% (n = 118). ARIA-H was the more frequently observed subtype, with an overall incidence of 22.4% (n = 105), while ARIA-E was identified in 12.2% of patients (n=57). Sixty-one cases had ARIA-H without ARIA-E (ARIA-H only = 13.0% of all patients); 13 cases had ARIA-E without ARIA-H (ARIA-E only = 2.8% of all patients); concurrent ARIA-H and ARIA-E (ARIA-Mixed) occurred in 44 cases (9.4% of all patients). ARIA events typically emerged early in the treatment course (Figure 1B), with ARIA-H emerging after an average of 8.6 infusions and ARIA-E after 6.4 infusions.

Most ARIA cases were detected on routine surveillance scans. Most ARIA-H events were radiologically mild (83.8%), followed by moderate (10.5%) or severe (5.7%) (Figure 1C). For ARIA-E, the majority were mild (59.6%), followed by moderate (38.6%), and only one case was severe when first discovered (1.8%) (Figure 1D). Twenty-three patients (19.5% of ARIA cases) with ARIA were symptomatic; the overall rate of symptomatic ARIA for this sample was 4.5%. Symptomatic ARIA rates were 6.6% of ARIA-H only cases, 30.8% of ARIA-E only cases, and 31.8% of ARIA-Mixed cases. The vast majority of symptomatic patients experienced mild symptoms except for one case of ARIA-E only, who experienced moderate symptoms of right leg weakness. Details on the demographic and clinical characteristics of patients with ARIA are presented in Table 2.

**Table 2.** Demographic and clinical information of the patients developing ARIA during lecanemab treatment.

|  |  | ARIA-H only (N=61) | ARIA-E only (N=13) | ARIA-Mixed (N=44) |
| --- | --- | --- | --- | --- |
| Variable |  | Mean ± SD or % | Mean ± SD or % | Mean ± SD or % |
| **Age** |  | 73.3 ± 6.9 | 67.4 ± 7.6 | 72.1 ± 7.9 |
| **Sex** | Female | 59.0% | 69.2% | 61.4% |
|  | Male | 41.0% | 30.8% | 38.6% |
| **Race** | White | 91.8% | 92.3% | 93.2% |
|  | Black | 1.6% | - | 2.3% |
|  | Asian | 3.3% | - | 2.3% |
|  | Other/unavailable | 3.3% | 7.7% | 2.3% |
| **Ethnicity** | Non-Hispanic | 90.2% | 76.9% | 88.6% |
|  | Hispanic | 6.6% | - | 2.3% |
|  | Unavailable | 3.3% | 23.1% | 9.1% |
| **Handedness** | Right | 75.4% | 69.2% | 90.9% |
|  | Left | 19.7% | 23.1% | 4.5% |
|  | Other | 4.9% | 7.7% | 4.5% |
| **APOE e4 status** | Non-carrier | 27.9% | 38.5% | 25.0% |
|  | Heterozygote | 62.3% | 46.2% | 52.3% |
|  | Homozygote | 9.8% | 15.4% | 22.7% |
| **FAQ Total Score** |  | 8.9 ± 5.5 | 8.8 ± 6.3 | 8.7 ± 6.4 |
| **QDRS Total Score** |  | 4.6 ± 2.3 | 4.4 ± 2.4 | 4.6 ± 2.4 |
| **MMSE Total Score** |  | 24.9 ± 3.6 | 24.3 ± 2.3 | 25.8 ± 2.3 |
| **Smoking** | Never | 63.9% | 61.5% | 65.9% |
|  | Former | 32.8% | 30.8% | 65.9% |
|  | Current | 3.3% | 7.7% | 4.5% |
| **Infusion number at the time of ARIA** |  | 22.9 ± 10.3<br>(range = 1-42) | 17.6 ± 10.4<br>(range 1-30) | 11.2 ± 9.0<br>(range = 2-33) |
| **Radiologic severity of ARIA** | Mild | 83.8% | 59.6% | 54.5% |
|  | Moderate | 10.5% | 38.6% | 43.2% |
|  | Severe | 5.7% | 1.8% | 2.3% |
| **Rate of symptomatic ARIA** |  | 6.6% (N=4) | 30.8% (N=4) | 31.8% (N=14) |
| **Clinical severity of symptoms** | Mild | 4 | 3 | 14 |
|  | Moderate | 0 | 1 | 0 |
|  | Severe | 0 | 0 | 0 |

### Cumulative incidence curves

Cumulative incidence curves were generated using the Kaplan-Meier estimator. To visualize the distribution of event severity, individual ARIA occurrences were plotted as points overlaid on the incidence curves, with colors indicating radiologic severity (Figure 2). The time-to-event distributions differed significantly between ARIA subtypes (log-rank test, p=0.012), with ARIA-H occurring more frequently and earlier than ARIA-E during the treatment course.

**Figure 2.**
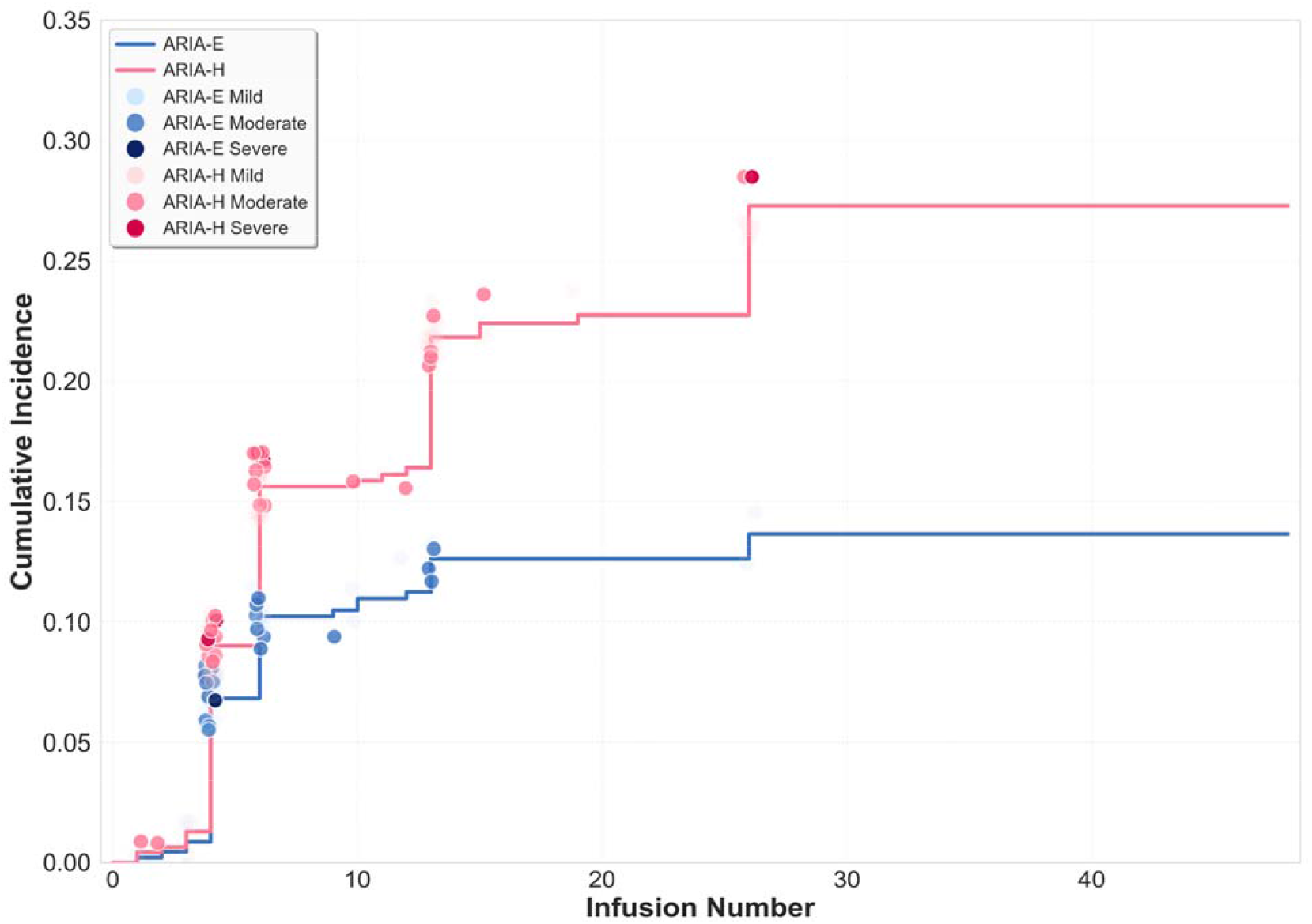
Cumulative Incidence of ARIA Events by Severity and Type. Individual events are overlaid and color-coded by radiologic severity. The time-to-event distributions for the two ARIA subtypes were significantly different (log-rank test, p=0.012)

### Regional Analyses of ARIA

The regional distribution of ARIA revealed distinct patterns for ARIA-E and ARIA-H (Figure 3, Supplementary Table 2). ARIA-H demonstrated elevated prevalence (p_FDR_< 0.05, indicated by asterisks) in frontal, left temporal, and left occipital regions. In contrast, ARIA-E was most prevalent in occipital regions. No left–right hemispheric asymmetry was detected for either ARIA-H (p=0.121) or ARIA-E (p=0.674).

**Figure 3.**
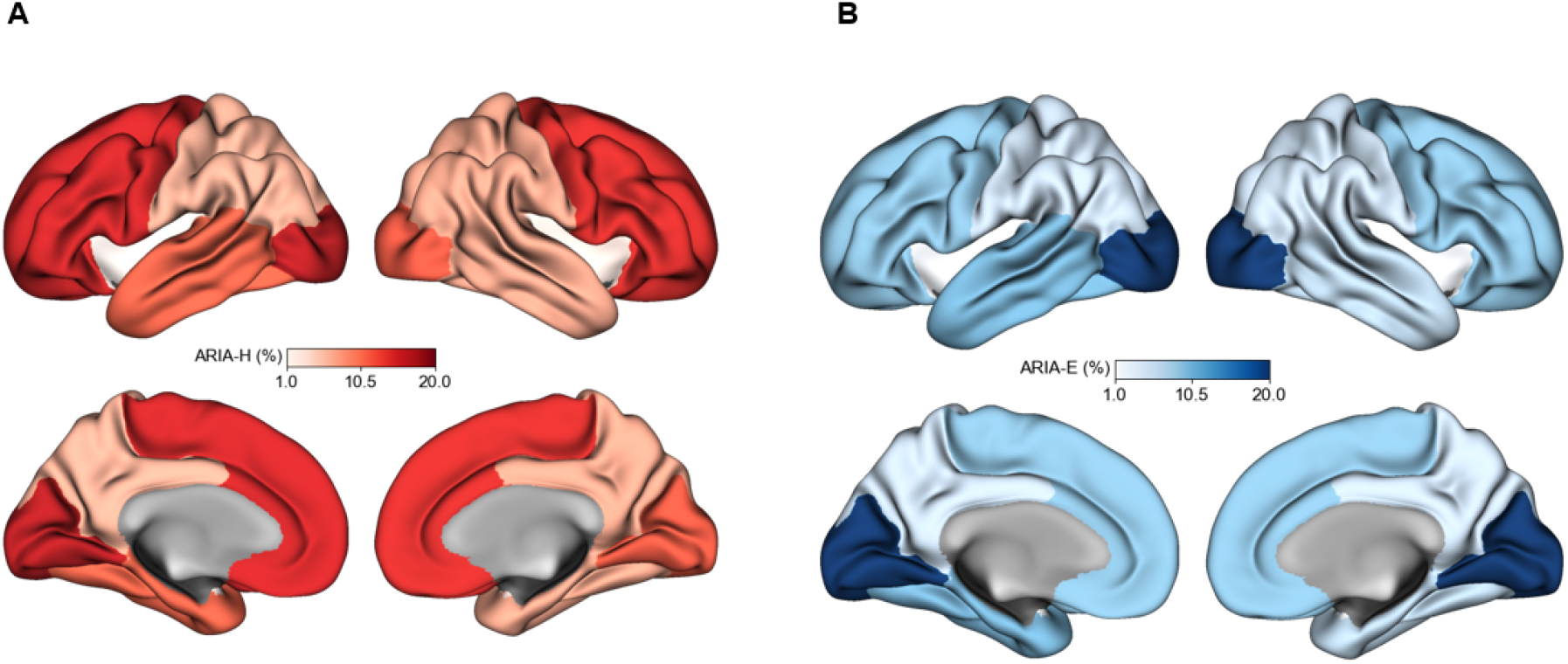
Regional cortical prevalence of ARIA-H (A) and ARIA-E (B). Darker colors indicate higher prevalence within each ARIA type.

### Natural history of ARIA

The joint longitudinal-survival model analysis revealed divergent patterns of natural history for ARIA-H and ARIA-E (Figure 4). Among patients who developed ARIA-H, severity remained stable with no increase in 69.6%, while worsening occurred in 30.4%. No cases showed complete radiologic resolution at the time of the most recent scan. Progression from mild to moderate or higher occurred in 15.1% of mild baseline cases, while progression from moderate to severe occurred in 54.5% of moderate baseline cases. In cases that worsened, the mean time to first increase was 64.3 ± 52.1 days (range 23–206), with similar timing across starting severities: 65.2 ± 54.9 days for mild, 65.9 ± 50.7 days for moderate, and 48.5 ± 13.5 days for severe.

**Figure 4.**
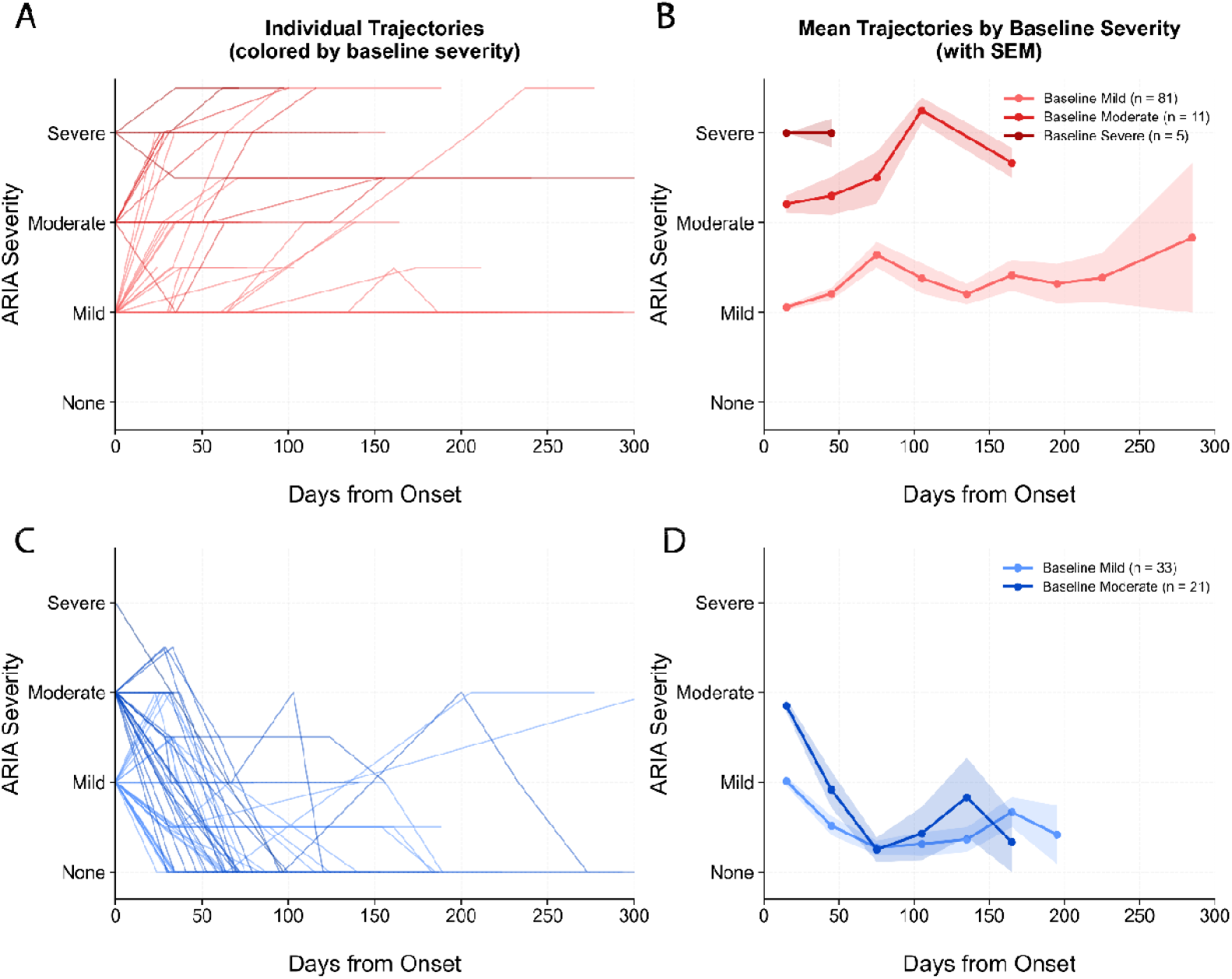
Natural history of ARIA-H and ARIA-E following initial radiologic detection. **(A)** Individual longitudinal severity trajectories for patients with ARIA-H colored by baseline severity at first detection. **(B)** Mean severity trajectories (± standard error of the mean) for ARIA-H, stratified by baseline severity category. **(C)** Individual longitudinal trajectories for patients with ARIA-E colored by baseline severity. **(D)** Mean severity trajectories (± SEM) for ARIA-E, stratified by baseline severity.

Among patients who developed ARIA-E, the natural history was generally favorable, with complete radiologic resolution in 80.4% overall. Severity remained stable with no increase in 10.7%, and worsening occurred in 8.9%. Transition from mild to moderate or higher occurred in 17.6% of mild baseline cases, while no moderate baseline cases progressed to severe. The overall mean time to radiologic resolution was 75.6 ± 51.0 days (range 24–273). By baseline severity, mean time to resolution was 72.8 ± 49.9 days for mild cases, 78.7 ± 53.6 days for moderate cases, and 97.0 days for the single severe case. In the small subset of non-resolved cases with no increase, the mean stability duration was 114.3 ± 91.0 days (range 0–277). In cases that worsened, the mean time to first worsening was 62.7 ± 74.7 days (range 20–314). Since our protocol includes monthly monitoring until resolution, the small proportion of non-resolved cases and ongoing follow-up times likely reflect active observation at the time of data extraction rather than permanent persistence in most instances.

### Univariate Predictors of ARIA

The incidence rates of ARIA-H and ARIA-E were correlated (r=0.423, p_FDR_<0.0001). In univariate analyses of baseline predictors of ARIA-H in the participant characteristic domain, fewer APOE ε3 alleles (r=-0.119, p=0.010, p_FDR_=0.049) and more APOE ε4 alleles (r=+0.128, p=0.006, p_FDR_=0.049) were associated with higher ARIA-H. In the AD biomarker domain, lower CSF Aβ42 demonstrated a robust inverse association (r=−0.202, p=0.010, p_FDR_=0.035). In the laboratory/vitals/imaging domain, higher baseline microhemorrhage count emerged as a strong predictor (r=+0.164, p<0.001, p_FDR_=0.005), while serum sodium (r=0.103, p=0.028) exhibited a nominal association that did not survive correction (p_FDR_=0.397 and 0.408, respectively). See Figure 5 for results.

**Figure 5.**
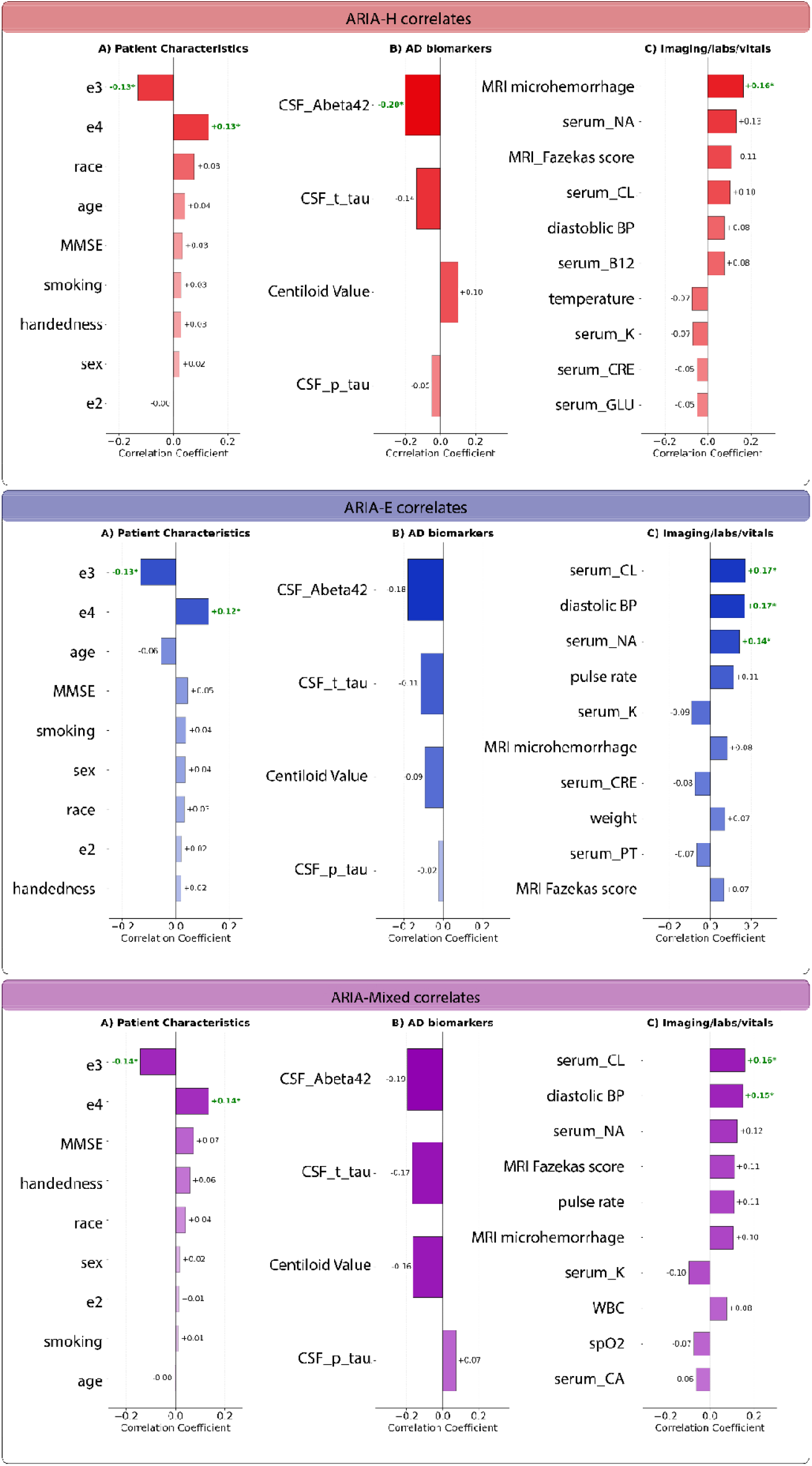
Baseline predictors of ARIA-H (top panel), ARIA-E (middle panel), and ARIA-Mixed (bottom panel) are shown through horizontal bar plots, using a graduated color intensity scheme where darker hues indicate stronger associations and lighter hues represent weaker correlations. Correlations that survive the FDR correction are shown in green.

In univariate analyses of baseline predictors of ARIA-E, within the participant characteristic domain, fewer APOE ε3 alleles (r=−0.128, p=0.005, p_FDR_=0.0493) and more APOE ε4 alleles (r=+0.123, p=0.008, p_FDR_=0.0494) reached nominal significance, whereas lower CSF Aβ42 was nominally associated in the AD biomarker domain (r=−0.195, p=0.015) but only showed a trend-level effect with multiple-comparison correction (p_FDR_=0.0731). In contrast, the laboratory/vitals/imaging domain yielded robust predictors that survived FDR correction: higher diastolic blood pressure (r=+0.169, p<0.001, p_FDR_=0.004), higher serum chloride (r=+0.172, p=0.0002, p_FDR_=0.0245), and higher serum sodium (r=+0.144, p=0.002, p_FDR_=0.025).

Among the subset of patients who developed both ARIA-Mixed, univariate analyses across baseline variables revealed several FDR-significant associations. These included more ε4 alleles (r=+0.135, p_FDR_=0.003), higher serum chloride (r=+0.159, p_FDR_<0.001) and higher diastolic blood pressure (r=+0.148, p_FDR_=0.002).

Furthermore, in a subanalysis of patients who developed symptomatic ARIA of any type, elevated diastolic blood pressure was strongly associated with symptomatic ARIA (r=0.23, p_FDR_<0.001).

### Survival Analysis for ARIA Risk Stratification

To identify clinically actionable thresholds for risk stratification, we performed survival analysis with maximally selected rank statistics on variables that showed significant univariate associations with ARIA to determine cutpoints (Table 3) and Kaplan-Meier curves for each risk factor (Supplementary Figure 1). For ARIA-H, patients with ≥1 microhemorrhage at baseline had a substantially increased risk compared with those without any microhemorrhages, while low CSF Aβ42 levels emerged as a particularly strong risk factor and APOE ε4 carriers (≥1 allele) exhibited a higher risk than non-carriers. For ARIA-E, elevated diastolic blood pressure was the strongest predictor, with values >75 mmHg associated with markedly increased risk, alongside elevated serum chloride (>105 mmol/L), elevated serum sodium (>141 mmol/L), and APOE ε4 carrier status (≥1 allele). For ARIA-Mixed (concurrent ARIA-H and ARIA-E), the same three factors—diastolic blood pressure >75 mmHg, serum chloride >105 mmol/L, and APOE ε4 carrier status (≥1 allele)—were associated with increased risk.

**Table 3.** Optimal cutpoints and hazard ratios for baseline predictors of ARIA in patients receiving lecanemab.

| ARIA Type | Variable | Cutpoint for Higher Risk | Hazard Ratio (95% CI) | p<sub>FDR</sub>-value |
| --- | --- | --- | --- | --- |
| ARIA-H | APOE ε4 Allele count | $\geq 1$ | 1.77 (1.14–2.75) | 0.003 |
| ARIA-H | CSF Aβ42 | $\leq 683.8$ | 6.39 (2.26–18.04) | $< 0.001$ |
| ARIA-H | Microhemorrhage count | $\geq 1$ | 2.55 (1.55–4.20) | $< 0.001$ |
| ARIA-E | APOE ε4 Allele count | $\geq 1$ | 2.51 (1.32–4.77) | 0.002 |
| ARIA-E | Diastolic Blood Pressure | $> 75.0$ | 3.12 (1.79–5.44) | $< 0.001$ |
| ARIA-E | Serum Chloride | $> 105$ | 2.79 (1.63–4.78) | $< 0.001$ |
| ARIA-E | Serum Sodium | $> 141.0$ | 1.70 (0.99–2.91) | 0.001 |
| ARIA-Mixed | APOE ε4 Allele count | $\geq 1$ | 2.87 (1.45–5.69) | 0.003 |
| ARIA-Mixed | Diastolic Blood Pressure | $> 75.0$ | 2.60 (1.43–4.74) | 0.002 |
| ARIA-Mixed | Serum Chloride | $> 105$ | 3.92 (2.11–7.27) | $< 0.001$ |

## Discussion

This real-world characterization of ARIA in patients receiving lecanemab provides a granular, clinically actionable understanding of the incidence, localization, natural history, and risk stratification of this potentially treatment-limiting adverse event. By leveraging comprehensive electronic health record data from a large, real-world clinical cohort (n=468 patients), we extend findings beyond the pivotal Clarity AD trial.^1^

The overall incidence of any ARIA in 25.2% of lecanemab-treated patients (ARIA-H 22.4%, ARIA-E 12.2%) is higher than that in the phase 3 Clarity AD trial, where any ARIA occurred in 21.3% of the lecanemab 10 mg/kg arm (ARIA-H 17.3%, ARIA-E in 12.6%).^1^ This is likely because our program employs 3-Tesla MRI scanners; higher field-strength imaging considerably increases sensitivity for ARIA-H.^30^

The differences in regional distribution of the two types of ARIA may reflect differential vascular amyloid burden, blood-brain barrier permeability, or local inflammatory responses across cortical regions, and warrant further investigation with advanced MRI modalities such as quantitative susceptibility mapping and a comparison with the localization of amyloid clearance.

The natural history of ARIA-E and ARIA-H is distinct. ARIA-H exhibited a chronic and persistent course. Radiologic resolution did not occur, and microhemorrhages tended to accumulate over time. In the absence of ARIA-E, ARIA-H was rarely symptomatic. In contrast, ARIA-E demonstrated a transient trajectory, with radiologic resolution occurring in most cases, independent of initial severity. This pattern supports current management guidelines on radiologic and clinical monitoring.^20^

In examining risk factors for ARIA, we confirmed that the presence of at least one APOE ε4 allele confers increased risk for both ARIA-E and ARIA-H. Furthermore, we showed that the presence of any microhemorrhages at baseline (≥1 microhemorrhage) and very low CSF Aβ42 (≤683.8 pg/mL) are potent predictors of ARIA-H. Lower baseline CSF Aβ42 was associated with increased risk of both ARIA-E and ARIA-H, although the association with ARIA-E did not survive correction for multiple comparisons. These findings are directionally consistent with the GRADUATE I/II trials of gantenerumab, where lower CSF Aβ42 was a strong independent predictor of ARIA-E as well as ARIA-E accompanied by concurrent new ARIA-H.^19^ We did not see this effect when amyloid PET Centiloids were analyzed, possibly due to the small sample size. Very low CSF Aβ42 reflects a more severe overall amyloid burden that often includes extensive deposition of amyloid in cerebral vessel walls (cerebral amyloid angiopathy, CAA).^25,31,32^ When the antibody binds to this vascular amyloid, it provokes local inflammation and vessel-wall fragility.^25,31,33^

Furthermore, higher baseline serum sodium and chloride levels are associated with increased risk of ARIA in lecanemab-treated patients, particularly ARIA-E. Although the observed values remained largely within or slightly above the conventional reference range, these findings align with emerging evidence that even moderate dietary salt excess promotes cerebrovascular fragility.^34^ Excessive salt, associated with hypertension,^35,36^ exacerbates vascular inflammation, impairs endothelial function, and restricts cerebral blood flow through pro-inflammatory mechanisms.^34,37^ In the context of anti-amyloid immunotherapy, pre-existing endothelial dysfunction and heightened vascular fragility from chronic high-normal sodium/chloride exposure may amplify the vasculotoxic effects of antibody-bound fibrillar amyloid in vessel walls, particularly in the setting of CAA.

Elevated baseline diastolic blood pressure (DBP >75 mmHg) emerged as a strong, modifiable predictor of ARIA-E in our cohort, and was strongly associated with symptomatic ARIA. Chronic hypertension, even at modest levels, impairs cerebral endothelial function, reduces vascular compliance, and disrupts pericyte-mediated blood-brain barrier.^38^ This pathophysiology is further complicated by evidence that hypertension may also impair the vascular clearance of amyloid-β, thereby facilitating its accumulation within cerebral vessels and parenchyma, a process that further heightens ARIA risk.^38,39^ In the context of lecanemab treatment, anti-Aβ antibodies bind to vascular amyloid and incite focal inflammation. Vessels already compromised by elevated diastolic pressure are less resilient to this insult, leading to increased vascular permeability and culminating in vasogenic edema characteristic of ARIA-E.

Our study has several limitations that should be considered when interpreting the results. First, some patients remain relatively early in their treatment course and may not have yet developed ARIA. While this could affect incidence estimates, the average of 20 infusions (which is about 10 months since treatment onset) and the minimum of 90 days into the treatment plan place the cohort beyond the typical peak period for ARIA development. Second, uniform analysis of amyloid data was limited, as patients in most cases underwent either lumbar puncture or amyloid PET imaging to determine amyloid status. Third, participants were recruited from a tertiary academic medical program, which limits the generalizability of incidence rates and risk factor thresholds to community practice settings.

This real-world analysis delivers a comprehensive characterization of ARIA during lecanemab treatment, detailing its incidence, distinct natural history, and regional topography. It reinforces established biomarkers while uncovering novel, modifiable predictors that may offer emerging mechanistic clues into ARIA pathogenesis. By integrating this multifaceted biological profile, we advance toward personalized risk stratification, enabling more tailored monitoring and potential preemptive management. To translate these findings into clinical care, future work should validate the identified risk thresholds in external cohorts, develop integrated multivariable risk scores, and investigate the underlying pathophysiology of novel associations. These findings also underscore the critical importance of systematically collecting rich, multidimensional data in large, accessible real-world registries to further refine the safety of amyloid-targeting therapies.

## Data Availability

The IRB protocols do not permit the sharing of the data outside the Mass General Brigham healthcare system.

## Declarations of interests

JRD has served on a scientific advisory board for I-MAB Biopharma. MWA is a co-founder of Aromha, Inc, and a consultant for Merck, TikTwo Therapeutics, Sudo Therapeutics, and has received in kind support from Lilly and IFF. SEA reports grants from Abbvie, AC Immune, Alzheimer’s Association, Athira, Challenger Foundation, Challenger Foundation, Ionis Pharmaceuticals, Janssen Pharmaceuticals, John Sperling Foundation, NIH, Novartis, Seer Biosciences, Venture Well, Gatehouse Bio, Eli Lilly/Fortrea, and SuperFluid Dx; consulting fees from Allyx Therapeutics, BioVie, Bob’s Last Marathon, Merck, Jocasta Neuroscience, Sage Therapeutics, Sanofi, and Vandria; and payment for expert testimony for Foster & Eldredge and ProSelect Insurance Co. DC is currently employed by Alnylam Therapeutics; this work was completed prior to her employment at Alnylam Therapeutics. SNG has served as a consultant for CervoMed, WaveBreak, Reachmr, Guidepoint Global, and Clearview and has served on a Steering Committee for Hillhurst Biopharmaceuticals and a Data Safety Monitoring Board for Ono. RYH has served as a scientific advisor to Vysioneer and has provided consulting for Telix, Meric-Cro, and Servier. ADL has served as a consultant for Acadia, Neurona Therapeutics, and UCB, and the institution of ADL has received research grant support from Neurona Therapeutics and Sage Therapeutics. BCD has served as a consultant for Acadia, Arkuda, Biogen, Cervomed, Eisai, Lantheus, Lilly, Merck, Novo Nordisk, Quanterix, and receives publishing royalties from Cambridge University Press, Elsevier, Oxford University Press, and Up To Date. JNF has received lecture fees from Life Molecular Imaging. GAM has received salary support from Eisai Inc. and Eli Lilly and Company for serving as a site principal investigator for clinical trials and has received payments for serving as a consultant for Ono Pharma USA, Inc. DJS is a founding director of Prothena Biosciences and ad hoc consultant to Roche and Eisai. TG-I declares grants or contracts from the National Institutes of Health, CureAlz, and MassCATS; payment or honoraria for lectures from Fundacion Tatiana de Guzman el Bueno; support for attending meetings and travel from CIBERNED (The Centre for Networked Biomedical Research in Neurodegenerative Diseases) and CIBER, University Complutense of Madrid, and Spanish Society of Neurology; participation on data safety monitoring boards or advisory boards for Periscope-ALZ trial, MindImmune, Mount Sinai ADRC (Alzheimer Disease Research Center), ACAD (Asian Cohort for Alzheimer’s disease) program at the University of Pennsylvania and Achucarro Basque Center for Neuroscience.

## Contributors

NR, JRD, and BCD contributed to the conceptualization of the project. NR, JRD, YH, JNF, LRG, SD, and BCD contributed to the data curation. NR and BCD contributed to the formal data analysis. NR and BCD contributed to the funding acquisition. NR, JRD, JNF, YH, YK, LRG, AL, HS, SA, MA, SRB, AB, PMB, DC, MD, ME, RG, SG, AG, MA, RYH, IK, SL, MM, SM, NP, SR, EH, JR, ASP, AS, AS, JY, GY, JC, KRD, MGE, SG, TGI, GAM, SMM, DS, HY, WWY, SD, BCD contributed to the investigation. NR, JRD, JNF, YH, SD, LRG, and BCD contributed to the methodology. NR and BCD contributed to the supervision of the project. NR, JRD, JNF, and BCD contributed to the validation of the data. NR and YK contributed to the visualization of the data. NR, JRD, SD, and BCD contributed to writing the original draft of the manuscript. All authors contributed to revising and editing the manuscript. All authors had the ability to access the data and had final responsibility for the decision to submit the manuscript for publication. NR, JRD, and BCD accessed and verified the data.

## Acknowledgements

The work described in this manuscript would not have been possible without the invaluable contributions of many MGB clinicians who are part of the MGB Alzheimer Therapeutics Program. The authors would like to specifically acknowledge the contributions of Yvette Bain, Katerine Betanco, and Abigail Shaughnessy. The authors also acknowledge the dedication of the infusion center, neurology, and radiology staff caring for patients treated with anti-amyloid therapy.

## Notes

### Author Declarations

This retrospective cohort study was conducted under MGB Institutional Review Board-approved protocols (2005P000860 and 2025P001042).

