## Supplemental materials for "Real-World Characterization of Amyloid-Related Imaging Abnormalities (ARIA) in Lecanemab Treatment at an Academic Health System"

### **Supplementary Methods**

**MRI monitoring protocol in the MGB APT**

ARIA monitoring was performed with a specifically developed MRI protocol modeled on ADNI sequences consisting of a three-dimensional (3D) T1-weighted sequence, a 3D FLAIR sequence, a GRE sequence, an Susceptibility-Weighted Imaging (SWI) sequence, and a Diffusion-Weighted Imaging (DWI) sequence. This MRI protocol was performed at baseline (pre-treatment) and at all routine monitoring MRIs. Neuroradiologists perform visual reads of the sequences following a standard ARIA reporting template. The detection of ARIA-E was performed with the FLAIR sequence to identify new parenchymal edema or sulcal effusions compared to baseline images. The detection of ARIA-H was performed with the GRE and the SWI sequence—each reported separately—to identify new microhemorrhages or superficial siderosis compared to baseline images. Here we focus our analysis on the GRE results.

Standard ARIA monitoring for lecanemab followed appropriate use recommendations, with scheduled MRI scans at baseline and prior to the 5th, 7th, and 14th infusions. For additional safety monitoring, the MGB ATP also performs monitoring prior to the 20th lecanemab infusion for patients with a history of ARIA requiring a pause in treatment due to ARIA severity and prior to the 27th lecanemab infusion for all patients. In addition to the standard ARIA monitoring schedule, an urgent MRI could be ordered by a clinician at any point in the treatment course due to a clinical concern for possible ARIA. Subsequently, based on FDA guidance, an additional surveillance MRI was added prior to the third infusion (data not included since that was a relatively recent development). If ARIA is detected, our standard protocol is to monitor it with monthly MRI scans until stabilization (ARIA-H) or resolution (ARIA-E).

Severity of ARIA-H is determined primarily by the number of new incident lesions. New microhemorrhages are generally classified as mild when ≤4, moderate when 5–9, and severe when ≥10. Superficial siderosis is graded as mild with one focal area, moderate with two focal areas, and severe with more than two focal areas or widespread involvement.^1^ Severity of ARIA-E is determined by the extent and distribution of the findings: mild ARIA-E is characterized by abnormalities measuring less than 5 cm in largest dimension and confined to a single cortical or subcortical location; moderate ARIA-E involves lesions 5 to 10 cm in size or multiple discrete sites; and severe ARIA-E is defined by lesions exceeding 10 cm, involvement of an entire lobe.^1^

**Electronic health record data extraction and data harmonization**

Multimodal patient data were extracted retrospectively from the Mass General Brigham Alzheimer’s Therapeutic Program template within the electronic health records for all individuals who initiated lecanemab treatment. The timeframe of the chart review was from the beginning of the Alzheimer Therapeutics Program, September 1, 2023, through the end of this analysis on November 7, 2025. We employed an automated pipeline, supplemented by an extensive manual data extraction, to minimize missing data for generating the required tabular data. We developed an AI-assisted natural language processing (NLP) pipeline to extract structured information from MGB ATP clinical notes using an on-premise, secure, IRB-approved (IRB Protocol number 2025P001042) installation of GPT. The workflow combines targeted preprocessing, GPT-based parsing, and post–quality control. Key sections were isolated with regular expressions and then parsed using GPT. Extraction templates were informed by clinician-designed Smart Phrases—capturing structured information on cognitive evaluations, AD biomarkers, MRI findings, and APOE genotypes—used in ATP clinic documentation. The model produced structured JSON outputs, which were flattened into tabular datasets and standardized through post-processing. Numeric fields such as PET SUVRs and MoCA/MMSE subscores were validated for internal consistency, and all outputs were linked to visit-level metadata. This pipeline enables consistent, reproducible extraction of structured variables from the clinical texts.

All baseline measures, including laboratory results, were collected within one year prior to the first lecanemab dose, with most obtained within three months. Variables included in this analysis are divided into three main categories: patient characteristics, AD-specific biomarkers, and imaging/labs/vitals, as shown in Supplementary Table 1. Although the vast majority of demographic and clinical variables used in the analysis to predict ARIA were available from most of the sample, baseline CSF or PET Centiloid values were available on only about a third of the sample each.

**Conversion of MoCA to MMSE Scores**
To maximize the use of cognitive assessment data and ensure consistency across the cohort, we converted Montreal Cognitive Assessment (MoCA) scores to their Mini-Mental State Examination (MMSE) equivalents. This conversion was performed using a validated mapping table, where each integer MoCA total score (0-30) was assigned its corresponding MMSE score based on established weighted mean values.^2^ The MoCA to MMSE conversion direction was specifically selected because previous validation studies have demonstrated it provides more accurate and reliable estimates compared to the reverse conversion, with narrower confidence intervals and better performance across the scoring range. The conversion was applied specifically for patients who had a recorded MoCA score but were missing an MMSE score, thereby imputing the missing MMSE data.

**Standardization of CSF Biomarker Assays**
Cerebrospinal fluid (CSF) biomarker measurements (Aβ42, t-tau, p-tau) were obtained using either Roche and ADMark® diagnostic assays. Previously, it has been shown that these assays have about 89% concordance.^3^ To harmonize measurements across assays for pooled analysis, we converted all ADMark®-derived values to Roche-equivalent values using robust distribution matching. Medians and interquartile ranges (IQRs) were computed for each biomarker separately within the ADMark® and Roche subsets of the dataset. For ADMark® samples, the transformation was applied as follows:

equivalent value = $\frac{ADMark® value - ADMark® median}{ADMark® IQR} \times Roche IQR + Roche median$

Roche values were directly assigned as equivalents without alteration. This approach is superior to traditional z-score normalization as it is more robust to outliers and skewed distributions, which are typical in CSF biomarker data. Based on Roche Laboratories, electrochemiluminescence immunoassays of AD pathology are defined by an Aβ42 concentration ≤ 834 pg/mL, a p-tau181 level > 21.6 pg/mL, and a t-tau > 238 pg/mL. Consequently, the p-tau181/Aβ42 ratio exceeds the diagnostic cut-off of >0.028, which has demonstrated 92% positive percent agreement with an abnormal amyloid PET scan, providing a robust indicator of underlying AD pathology.^4^

| Supplementary Table 1. Clinical and demographic variables analyzed in association with ARIA | | |
| --- | --- | --- |
| Domain | **Variable** | **Available data** |
| Patient characteristics | Age | 100% |
|  | Sex | 100% |
|  | Race | 97.4% |
|  | Handedness | 95.5% |
|  | Number of e2 alleles | 100% |
|  | Number of e3 alleles | 100% |
|  | Number of e4 alleles | 100% |
|  | Smoking (pack.years) | 83.5% |
|  | MMSE (harmonized) | 100% |
| AD biomarkers | CSF_Abeta42 (assay harmonized) | 34.6% |
|  | CSF_t_tau (assay harmonized) | 34.6% |
|  | CSF_p_tau (assay harmonized) | 34.6% |
|  | Amyloid PET Centiloid Values | 28.9% |
| Imaging/labs/vitals | Pulse rate | 95.7% |
|  | Temperature | 90.8% |
|  | Systolic blood pressure | 97.4% |
|  | Diastolic blood pressure | 97.9% |
|  | SPO2 | 94.2% |
|  | Height | 96.2% |
|  | Weight | 91.6% |
|  | MRI_microhemorrhage count (GRE) | 100% |
|  | MRI_Fazekas score | 96.4% |
|  | MRI.lacunar_infarcts | 98.1% |
|  | MRI.cortical_infarcts | 98.1% |
|  | serum_WBC | 99.1% |
|  | serum_RBC | 98.9% |
|  | serum_PLT | 99.1% |
|  | serum_INR | 95.3% |
|  | serum_PT | 87.8% |
|  | serum_PTT | 91.6% |
|  | serum_NA | 98.9% |
|  | serum_K | 98.9% |
|  | serum_CL | 98.5% |
|  | serum_CO2 | 98.5% |
|  | serum_BUN | 98.9% |
|  | serum_CRE | 99.1% |
|  | serum_GLU | 98.3% |
|  | Serum_CA | 98.5% |
|  | serum_ALKP | 97.6% |
|  | serum_SGOT | 96.6% |
|  | serum_SGPT | 96.6% |
|  | serum_B12 | 97.2% |
|  | serum_TSH | 98.9% |
| Table abbreviations: AD, Alzheimer disease; APOE, apolipoprotein E; B12, vitamin B12; BUN, blood urea nitrogen; CA, calcium; CL, chloride; CO₂, carbon dioxide; CRE, creatinine; CSF, cerebrospinal fluid; GLU, glucose; INR, international normalized ratio; K, potassium; MMSE, Mini-Mental State Examination; MRI, magnetic resonance imaging; NA, sodium; PET, positron emission tomography; PLT, platelets; PT, prothrombin time; PTT, partial thromboplastin time; RBC, red blood cells; SGOT, serum glutamic-oxaloacetic transaminase; SGPT, serum glutamic-pyruvic transaminase; SPO₂, peripheral capillary oxygen saturation; SUVR, standardized uptake value ratio; TSH, thyroid-stimulating hormone; WBC, white blood cells | | |

### **Supplementary Results**

Detailed regional prevalence of ARIA-H and ARIA-E is shown in Supplementary Table 2.

| Supplementary Table 2. Regional prevalence of ARIA-H and ARIA-E (FDR-corrected %) across brain structures. | | |
| --- | --- | --- |
| Region | ARIA-H % (FDR-corrected) | ARIA-E % (FDR-corrected) |
| right frontal | 14.5%* | 9.2% |
| left frontal | 15.0%* | 9.2% |
| right insular | 2.3% | 0.0% |
| left insular | 1.2% | 0.9% |
| right temporal | 6.9% | 6.4% |
| left temporal | 11.0%* | 10.1% |
| right parietal | 6.90% | 5.50% |
| left parietal | 6.40% | 5.50% |
| right occipital | 11.6%* | 27.5%* |
| left occipital | 16.2%* | 23.9%* |
| right brainstem | 0.6% | 0.0% |
| Left brainstem | 0.0% | 0.0% |
| right centrum semiovale | 0.0% | 0.0% |
| left centrum semiovale | 0.6% | 0.0% |
| right corpus callosum | 0.0% | 0.9% |
| left corpus callosum | 0.0% | 0.0% |
| right basal ganglia | 1.2% | 0.0% |
| left basal ganglia | 0.0% | 0.0% |
| right brainstem | 0.0% | 0.0% |
| left brainstem | 0.6% | 0.0% |
| right cerebellar | 1.2% | 0.0% |
| left cerebellar | 4.0% | 0.9% |

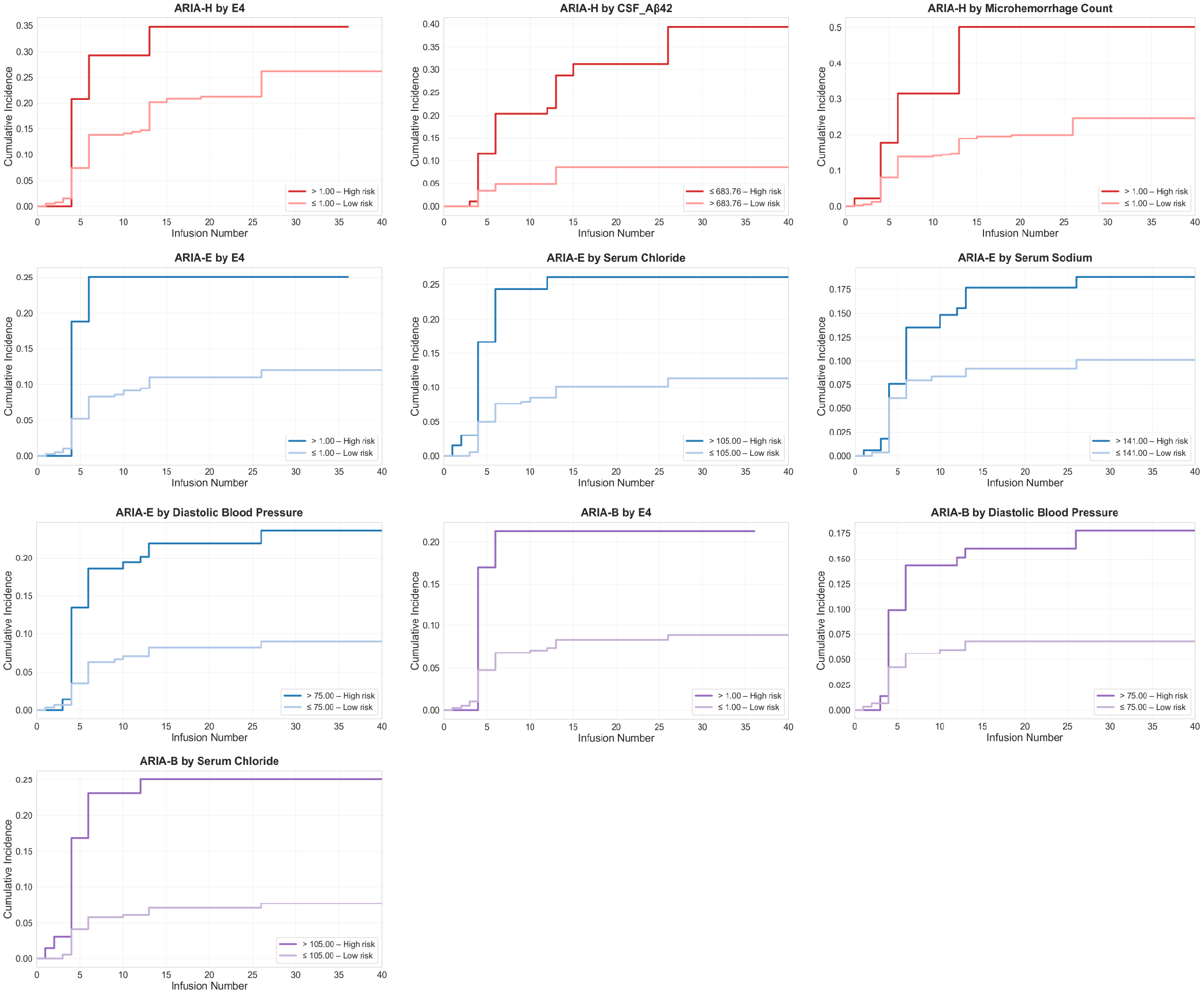

**Supplementary Figure 1.** Kaplan-Meier curves showing cumulative incidence of ARIA-H (red), ARIA-E (blue), and ARIA-mixed (purple) stratified by optimal data-driven cutpoints for key baseline risk factors, with global Benjamini-Hochberg FDR correction
